# Safety and efficacy of Pulmonary physiotherapy in hospitalized patients with severe COVID-19 pneumonia (PPTCOVID Study): A prospective, randomised, single-blind, controlled trial

**DOI:** 10.1101/2021.04.24.21255892

**Authors:** Mohammad Javaherian, Azadeh Shadmehr, Abbasali Keshtkar, Mohammad-Taghi Beigmohammadi, Narges Dabbaghipour, Aabis Syed, Behrouz Attarbashi Moghadam

## Abstract

**Background:** Pulmonary physiotherapy (PPT) is an important treatment in the management of patients with different types of pulmonary disorders. We aimed to evaluate safety and efficacy of PPT in hospitalized patients with severe COVID-19 pneumonia.

**Methods:** In this randomised, single-blind, controlled trial, we enrolled hospitalized, non-intubated patients (18 to 75 years with oxygen saturation (Spo2) in free-air breathing ≤90%) with COVID-19 pneumonia at a referral hospital. Participants were randomly assigned (1:1) to receive PPT (six sessions PPT with breathing exercises and airway clearance techniques) or basic care. The primary outcomes were venous blood O2 (pO2) and CO2 (pCO2) pressures, Spo2, and three-minute walking test (3MWT) that were assessed before and end of sixth session. Secondary outcomes included level of dyspnea, venous blood PH, one-month mortality, three-month mortality and short form-36 (SF-36) after one and three months. The assessor was blinded to the assignment. This trial is registered with ClinicalTrials.gov (NCT04357340).

**Findings:** In April-May 2020, 40 participants were randomly assigned to PPT or basic care groups. While at the end of intervention, pO2 (adjusted mean difference to baseline measure (AMD) 6.43 mmHg [95%CI 2.8, 10.07], P<0.01), Spo2 (AMD 4.43% [95%CI 2.04, 6.83], P=0.0011), and 3MTW (AMD 91.44 m [95%CI 68.88, 113.99], P<0.01) were higher in PPT group and basic care group, pCO2 was not improved (AMD -2.1 mmHg [95%CI-6.36, 2.21], P=0.33). Mortality rate was 20% (OR adjusted to baseline Spo2 .19 [95%CI .03, 1.30], P=.09) and 25% (OR adjusted to baseline Spo2 .16 [95%CI .26, 1.05], P=.056) lower in the PPT group at three-month at one-month and three-month, respectively. There were no significant differences in most SF-36 domains scores after one and three months. No serious adverse event was observed during PPT sessions.

**Conclusion:** Early PPT can be considered a safe and effective therapeutic choice for patients with severe COVID-19.

## Introduction

In late December 2019, the coronavirus infection pneumonia epidemic broke out and rapidly spread worldwide [1]. Now almost all countries are challenging with coronavirus disease 2019 (COVID-19). Severe Acute Respiratory Syndrome Coronavirus 2 (SARS-CoV-2) not only induced more than 194 million definite COVID-19 cases and four million mortality numbers yet, but it also caused a challenge in healthcare systems and the economic conditions of most countries [2].

Patients with COVID-19 can present influenza-like signs and symptoms like fever (89%), cough (68%), fatigue (38%), excessive pulmonary secretion (EPS) or sputum (34%), and breathlessness (19%). The disease can appear asymptomatic or mild lower respiratory tract illness through severe viral pneumonia with acute respiratory distress syndrome and even death [3]. Age> 60 years, some underlying noncommunicable diseases like diabetes, hypertension, cardiovascular, kidney, and lung diseases, cerebrovascular diseases, immunosuppression, obesity, cancer, and even mental disorders are reported as risk factors associated with severe COVID-19 and mortality [3].

Pulmonary physiotherapy (PPT) is a comprehensive therapeutic method that is aimed to improve patient’s respiratory symptoms, train effective cough, clear the airways, decrease symptoms of dyspnea, re-expansion pf atelectatic lung, provide short-term improvement in lung-thorax compliance, reduce the complications related to disease, minimize disability and finally improve the health-related quality of life (HRQOL) [4-6].

Up to now, some studies with different designs and levels of evidence have been suggested PPT strategies for hospitalized patients with various stages of COVID-19 pneumonia [1, 4, 7-13].

Almost all of their recommendations are based on expert opinion origin and reported data from other viral pneumonia diseases such as influenza. These recommendations seem to have not sufficient experimental evidence in connection with COVID-19. The original studies have focused on rehabilitative or physical therapy processes after the severe stage of COVID-19 more than the acute stage to recovery muscle wasting, aerobic deconditioning, and possibly happened disability due to prolonged immobility [14].

Although some researchers and expert clinicians suggested that early PPT can be tailored in the condition of acute respiratory disease to prevent further muscular and aerobic systems deterioration, helping the therapeutic process, decrease hospitalization duration, and facilitate future recovery in patients with COVID-19 [4, 7, 11, 12], but there are some researchers recommend that use of this treatment should be avoided for patients with COVID-19 especially in the severe or critical stage [3] except in some circumstance like the presence of a high amount of EPS [9]. Most of these researchers believe that this treatment can be unsafe and ineffective for patients with acute COVID-19. Although some researchers reported that early PPT can be safe and be associated with clinical improvement, prevention of potential complications due to underlying diseases and immobilisation, short length of stay and a decrease of mortality rate in patients with critical diseases [15-18], but there is no high-quality randomised controlled trial (RCT) evaluate the safety and efficacy of PPT in patients with severe COVID-19 after more than 18 months from SARS-Cov-2 spread in the world. This study was aimed to evaluate the safety and efficacy of PPT on venous blood gas (VBG) analysis, peripheral blood oxygen saturation (Spo2), three-minute walking test (3MWT), one-month and three-month HRQOL and mortality rate in non-intubated patients with severe COVID-19 pneumonia. The result of this study can guide the clinicians and future researchers to include PPT into the protocol of severe COVID-19 management or not. It was hypothesized that participants receive PPT demonstrate improvement in venous blood O2 & CO2 pressures (pO2 and pCO2, mmHg), Spo2, 3MWT, level of dyspnea, one-month and three-month HRQOL, and decrease of one-month and three-month mortality rates than participants in the control group.

## Material and methods

### Study design

This was a single-center, single-blind, prospective, two parallel-armed RCT comparing the efficacy of six sessions PPT + basic care with basic care alone in hospitalized patients with COVID-19 pneumonia. In April and May 2020, participants were recruited at COVID-19 intermediate care wards and intensive care units of Imam Khomeini Hospital Complex, Tehran, Iran. IKHC is known as the largest and one of the referral hospitals in Iran. Any related adverse events during three days were monitored and recorded by a data safety monitoring committee. Members of this committee were physicians of intermediate care wards and one medical ethics specialist. They were asked to follow the procedure of applied interventions on patients by researchers and possible occurred adverse events which may be related to PPT. This trial was approved by the Ethical Committee of Tehran University of Medical Sciences (IR□JTUMS.VCR.REC.1399.205). The authors had access to patients’ medical records until three months after recruitment.

### Participants

All participants were patients with COVID-19 pneumonia at a severe stage [3]. The inclusion criteria were individuals: 1) 18 to 75 years old; 2) with confirmed COVID-19 by positive reaction real-time polymerase chain reaction test and presence of ground-glass opacification in their chest computed tomography scan; 3) severe stage of COVID-19 pneumonia indicated by Spo2 < 90% after two-minute breathing the free air and respiratory rate > 30 breaths/min ;4) with full consciousness and oriented; 5) able to walk; 6) with O2 saturation < 90% after; 7) 100 ≤ PaO2/FiO2 ≤ 200 mmHg, and 8) able to read and write in Persian. The exclusion criteria included 1) presence of any type of musculoskeletal disorder that prohibits the patient from participating in the study; 2) history of intubation due to COVID-19 pneumonia; and 3) Any type of obvious clinically mental or cognitive impairment. Potentially eligible participants were screened by review the admitted patient’s medical records and a preliminary assessment. A signed informed consent form was obtained from all patients after explaining the study details to them.

Participants who met one of the following conditions were withdrawn/dropped out from the study: 1) Unable to complete at least three sessions; 2) Dissatisfaction to continue to the study for any reason; 3) Intubation or die during interventions; and; 5) Occurring unpredictable adverse events during applying interventions.

### Randomisation and masking

After consent was obtained, eligible individuals were assigned (1:1) to one of the treatment groups using the blocked-balanced randomisation method [19] (block size: 4): 1) the experimental group (received six sessions PPT in addition to basic care interventions) or 2) the control group (received basic care only). Randomization was done using an online system that provides difference blocks order based on a random number (https://www.sealedenvelope.com). The sealed envelopes were provided and stamped based on the order of randomised groups by a researcher (BAM) who had no role in the participants’ assessment and treatment.

Physiotherapists allocated participants based on the order of the envelopes. The assessor and statistical analysis team were masked to treatment allocation. The assessor had not also access to patient’s medical record. As PPT consists of some techniques that require the patient’s cooperation actively; therefore, participants masking was impossible.

### Procedures

The process of enrollment, recruitment and the baseline assessment was performed in a same day. Medical history and demographic data were collected from participants after recruitment. Patients underwent clinical evaluation at baseline, including VBG analysis, 3MWT, Spo2, and level of dyspnea. In order to VBG analysis, blood samples were taken from the upper extremity peripheral veins after two-minutes of breathing free air in fowler’s position by a nurse. Samples were immediately analyzed [20]. At this time, the patient’s Spo2 was assessed using an index finger pulse oximeter [21]. In the 3MWT, patients were asked to continue walking as their tolerance without portable oxygen receive using comfortable footwear. Heart rate and Spo2 were monitored and the test was stopped in the condition of Spo2 < 80%, patient’s inability to continue, dyspnea, chest or limb pain [22]. Rated perceived exertion (RPE) during exercises was evaluated using the Borg scale immediately after 3MWT [23]. The level of dyspnea was evaluated using the visual analog scale [24]. To assess the level of dyspnea, the patient was instructed that zero stands for no breath shortness and 10 shows maximum breath shortness. Patient was asked to mark his/her feeling of dyspnea during the past 24 hours on the paper [25]. The interventions were delivered over three days. The number of days and sessions for applying the intervention was determined after analyzing the mean of staying duration at intermediate care wards, where patients with severe COVID-19 were admitted. Based on the result of primary analysis, 78% of patients had admitted to intermediate care wards stayed for 3-4 days and they were then transferred to other care wards (normal care wards or intensive care units). Patients underwent a second assessment time at the end of day three (after the sixth session). Participants were also followed up to one month and three months to evaluate their survival rate (using electronic medical records) and HRQOL using telephone assessment of short form-36 (SF-36) questionnaire.

### Basic care group

Participants in the basic care group received usual medical care, including medication based on the national guideline of COVID-19 [26], oxygen therapy, possibly non-invasive ventilation intervention, nursing care and monitoring, and etc. They also had one session of breathing education with 40 minutes duration includes effective cough, if they felt EPS, diaphragmatic breathing, and upper chest muscles relaxation. They also received an incentive spirometer and instructions on how to use it. They also were advised to walk as tolerate, even using a portable oxygen cylinder. This educational session was held in one of three days after the first assessment and before the second assessment.

### Pulmonary physiotherapy group

In addition to basic care interventions, patients allocated to the PPT group received a totally 6 PPT sessions for three consecutive days (twice daily: one session in the morning and one session in the afternoon). PPT was started at the same day that patient was recruited. Patients received two sessions of PPT in that day. Two expert pulmonary physiotherapists assessed patients immediately before every session to select the techniques based on their findings. It is reported that about 34% of patients with COVID-19 pneumonia present signs and symptoms of EPS [3]. If the patient presented signs and symptoms of EPS presence like wheezing sound, and sputum production, airway clearance techniques (ACTs) were applied. These techniques included active cycle of breathing (ACBT), Autogenic Drainage, vibration, and postural drainage. If there was any contraindication for these techniques, only an effective cough technique was used. ACTs were continued until successful secretions clearance based on therapists’ assessment [27]. These patients and whom without EPS signs and symptoms then received Inspiratory Hold Technique (IHT) exercises. During these exercises, patients held their breath at the end of inspiration without using a Valsalva maneuver following by a relaxed exhalation. The physiotherapists emphasized diaphragmatic breathing while performing the IHT by setting their palmar surface of hand on patients’ costal margin bilaterally to facilitate abdominal wall motion. Also, patients were educated to relax their upper chest muscles during exercises [27]. This technique was performed ten times in three sets with 2 minutes rest between each set: 1) Set 1: three-second hold, six-second rest; 2) Set 2: six-second hold; 12-second rest; 3) Set 3: 10-second hold, 20-second rest. A summary of applied PPT techniques, contraindications, and criteria of holding the interventions, is presented in Fig 1.

**Fig 1.**
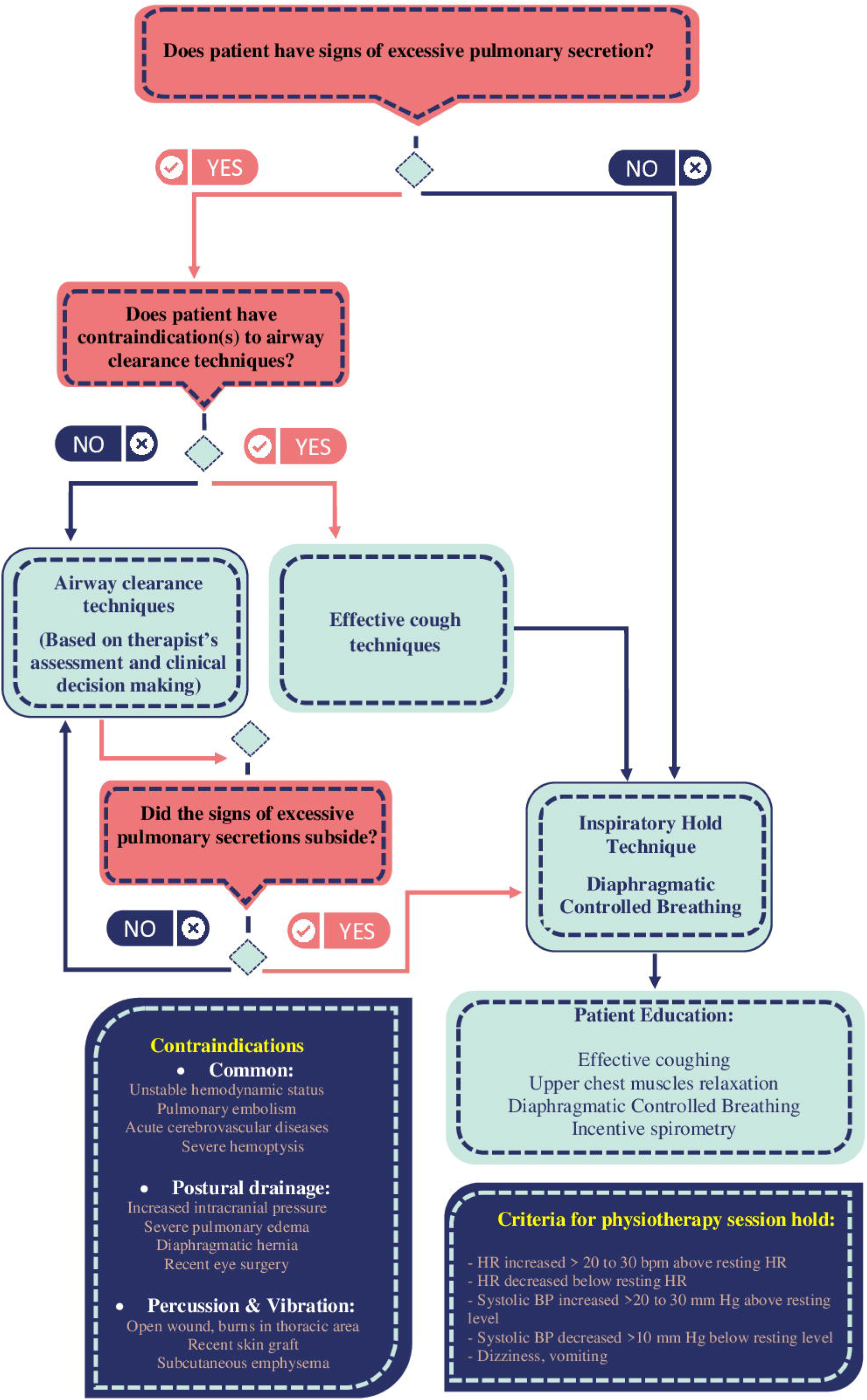
The protocol of applied pulmonary physiotherapy to patients with or without excessive pulmonary secretion.

Also, patients in this group underwent mild aerobic exercise by walking training after the third session. Two criteria indicated the end of walking training: 1) O2 saturation < 80% or 2) 11 < RPE < 13 based on the Borg’s scale [28]. The maximum walking training time was determined six minutes, and individuals could have rest intervals during this period.

### Outcome measures

The primary outcomes were pO2 and pCO2 from VBG analysis. Arterial blood gas analysis was not selected to measure the O2 & CO2 pressures due to two reasons; first, patients in the intermediate care have not artery line and also blood sampling by syringe is a painful procedure; second, it has been shown that VBG analysis can be considered as an acceptable alternative for arterial blood gas analysis in patients with pulmonary diseases [20]. Other primary outcome measures include Spo2 after 2 minutes breathing free air, and 3MWT distance.

The secondary outcomes included the level of dyspnea and RPE after 3MWT using Borg’s scale [15], PH of VBG, one-month and three-month related mortality rates and, HRQOL by the Persian version of SF-36 after one month and three months [29]. As recruited patients had severe disease and could not respond to SF-36 questions, we did not assess their HRQOL at the baseline.

### Statistical analysis

The sample size of this study was calculated based on minimal clinically importance difference (MCID) of Spo2. Considering standard deviation (SD) of 4.4%, MCID of 4%, two steps of measurements (before and after), a correlation between measurements of .6, type I error of .05, and 80% power, a sample size of 13 participants per group was calculated. [30, 31]. As participants were patients with severe COVID-19 pneumonia and the mortality rate seems to be high in this group, 50% loss to follow-up was estimated. Finally, 40 patients were considered as a suitable sample size.

Stata version 13. (Stata, College, Statin, Texas) was used to analyze the data. We reported Means ± SD and frequency counts (%) for continuous and categorical data, respectively. Data normality was checked for all continuous variables using the P-P plot, Q-Q plot, and Shapiro-Wilk test. Analysis of variance and covariance (ANOVA/ANCOVA) was used to determine the differences of all continuous data, with the baseline score included as a covariate (one factor, one covariate) [32, 33]. Also, we selected blood Spo2 as another covariate (one factor, two covariates) due to two reasons: 1) Spo2 can be considered one of the important factors to predict mortality in hospitalized patients with COVID-19 pneumonia [31, 33]. Second, the MD of Spo2 between groups was more than .2 × SD of all participants’ Spo2 [34]. The point estimates of effects were reported as MD with a 95% confidence interval (CI), standardized mean difference (SMD) with 95% CI analyzed by Cohen’s d. We considered .2 - .49, .5 - .79, .8 - 1.19 and, > 1.2 valuables as small, moderate, large, and very large SMD effects, respectively. To compare three analysis models and the effectiveness of cofounder/covariable impact on the results, partial eta2 effect size was provided. Change of partial eta^2^ > 10% between three analysis models was considered important. In the analysis of categorical data, intention-to-treat (ITT) analysis was utilized for all categorical data.

The number needed to treat (NNT) was calculated using an online calculator for all measures [35]. This system transforms Cohen’s d into NNT and therefore is difficult to interpret. Alpha ≤ .05 was considered a statistically significant level.

The study was registered at www.clinicaltrials.gov (ClinicalTrials.gov Identifier NCT04357340). The registration process was completed some days after recruitment and before primary completion. The reason why recruitment was not started after registration was our particular situation at that time in our center related to the pre-planned timetable for studies related to the COVID that the Research deputy of the hospital had determined. Therefore, we had to start study before having the registration number. The authors confirm that all ongoing and related trials for this drug/intervention are registered.

## Modifications to the study protocol

Level of dyspnea, RPE after the 3MWT, and PH from VBG analysis were considered as secondary outcome measurements, while they were identified as primary outcomes in registration. History of intubation due to COVID-19 pneumonia was considered as another exclusion criteria. Also, the survival rate and HRQOL assessment were evaluated after three months as second follow-up. The permissions for these amendments were obtained from the ethical committee. The details and reason for the protocol modifications are presented in S1.

## Results

In April and May 2020, 255 patients were screened, of whom 40 (age: 55.5 ± 13.7) met the criteria and were randomly allocated to PPT (n=20) or basic care (n=20) groups. From 20 allocated patients to the PPT group, one (5%) was intubated at the end of the second day and died after nine days. The reason for intubation was suddenly hemodynamic instability, which was diagnosed due to pulmonary embolism. Two patients (10%) in the control group were intubated on the third day due to pneumonia progress and died three and eight days later. These participants’ baseline data were included in the statistical analysis. After one and three months, four participants (10%, two in PPT and two in basic care groups) and five participants (12.5%, two in the PPT group and three in the control group) declined to participate in a telephone-based assessment of SF-36. We found all participant’s living status; therefore, no sample attrition occurred. Accordingly, ITT and per-protocol were the same for categorical data analysis. The trial profile is presented in Fig 2.

**Fig 2.**
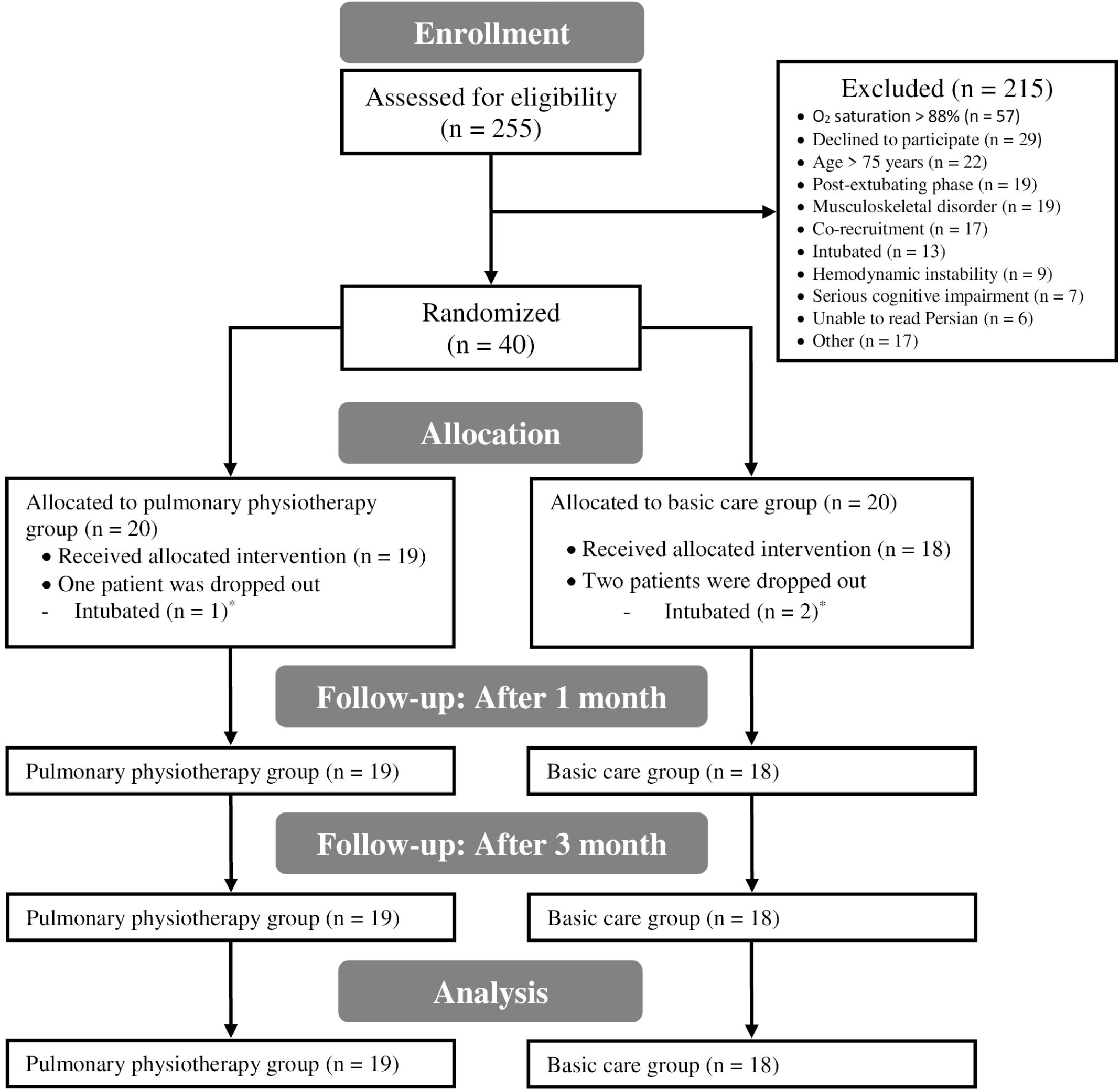
Trial profile.

The demographic and clinical characteristics of all participants are presented in Table 1.

**Table 1.** Demographics and clinical characteristics of participants.

|  | PPT group<br>(n=20) | Basic care<br>group<br>(n=20) |
| --- | --- | --- |
| Age (y) (Mean ± SD) | 54.4 ± 13.1 | 56.6 ± 14.4 |
| Sex |  |  |
| Male, n (%) | 13 (65) | 12 (60) |
| Female, n (%) | 7 (35) | 8 (40) |
| Height (cm) (Mean ± SD) | 172.4 ± 9.3 | 173.4 ± 7.4 |
| Weight (kg) (Mean ± SD) | 82.6 ± 11.8 | 79.8 ± 19.3 |
| BMI (kg/m<sup>2</sup>) (Mean ± SD) | 27.9 ± 4.3 | 26.2 ± 4.4 |
| Comorbidity, n (%) |  |  |
| Any comorbidity, n (%) | 10 (50) | 9 (45) |
| Diabetes mellitus, n (%) | 5 (25) | 2 (10) |
| Cardiovascular disease, n (%) | 4 (20) | 4 (20) |
| Pulmonary dysfunction (s), n (%) | 1 (5) | 1 (5) |
| Renal failure, n (%) | 0 (0) | 2 (10) |
| Others, n (%) | 2 (10) | 1 (5) |
| Smoker, n (%) | 3 (15) | 4 (20) |
| Occupational environmental exposure, n (%) <sup>a</sup> | 2 (10) | 3 (15) |
| Routine Ventilation Type<sup>b</sup> |  |  |
| Nasal, n (%) | 2 (10) | 3 (15) |
| Face mask, n (%) | 4 (20) | 3 (15) |
| Partial rebreathing mask, n (%) | 13 (65) | 12 (60) |
| CPAP, n (%) | 1 (5) | 2 (10) |
| Duration of Stay before enrolment (Mean ± SD) | 10.1 ± 11.5 | 9.5 ± 6.8 |
| Baseline O<sub>2</sub> saturation (%), Mean ± SD) | 83.4 ± 4.9 | 84.4 ± 3.9 |

PPT: Pulmonary physiotherapy; SD: Standard deviation; BMI: Body mass index; CPAP: Continuous positive airway pressure
<sup>a</sup> defines as patient's exposure to some materials like asbestos, arsenic, coal, etc., which can cause pulmonary disease.
<sup>b</sup> Defined as prescribed ventilation type for patient by physician which should be used most of the time.
\* All of the participant's characteristics are statistically similar between groups based on t-test and chi-square test.

In the crude model analysis, the mean pO2 (MD: 5.81 [1.51, 10.11], SDM: .9 [.21, 1.57]), Spo2 (MD: 3.99 [1.5, 6.47], SMD: 1.07 [.37, 1.76]), 3MWT distance (MD: 81.82 [47.93, 115.7], SMD: 1.61 [.86, 2.35]), level of dyspnea (MD: -2.08 [-2.96, -1.19], SMD: -1.57 [-2.31, -.82]), RPE after walking (MD: -4.04 [-5.76, -2.32], SMD: -1.56 [-2.3, -.82]) and, PH (MD: .06, [.01, .12], SMD: .74 [.71, 1.41]) were statistically significant different between groups in all analysis models (P ≤ .03, Table 2). Also, pCO2 was insignificantly decreased (MD: -1.68 [-6.8, 3.44], SMD: -.22 [-.43, .86]). In the ANCOVA with adjusting each variable to its baseline measure, the partial eta^2^ were improved > 10%. Considering baseline Spo2, partial eta^2^ were slightly reduced in all variables except level of dyspnea which was slightly increased (Table 2).

**Table 2.** Distribution of different outcomes according to two arms in addition to related effect sizes.

| Clinical Outcome | PPT group (n=19) |  | Basic care group (n=18) |  | F (P value) ** | MD [95 % CI] | SMD [95 % CI] | Partial eta<sup>2</sup> |
| --- | --- | --- | --- | --- | --- | --- | --- | --- |
|  | Baseline<sup>a</sup> | After<sup>a</sup> | Baseline<sup>a</sup> | After<sup>a</sup> |  |  |  |  |
| pO<sub>2</sub> (mmHg) | 29.4 ± 5.3 | 38.7 ± 5.3 | 30.6 ± 8.2 | 32.9 ± 7.5 | 7.5 (< .01)<sup>a</sup> | 5.81 [1.51, 10.11] | .9 [.21, 1.57] | .177 |
|  |  |  |  |  | 13 (< .01)<sup>b</sup> | 6.43 [2.8, 10.07] | 1.18 [.48, 1.88] | .276 |
|  |  |  |  |  | 12.04 (< .01)<sup>c</sup> | 6.39 [2.64, 10.14] | 1.15 [.45, 1.84] | .267 |
| pCO<sub>2</sub> (mmHg) | 40 ± 7.3 | 37 ± 7.2 | 39.3 ± 8 | 38.6 ± 8.1 | .44 (.51)<sup>a</sup> | -1.68 [-6.8, 3.44] | -.22 [-.43, .86] | .012 |
|  |  |  |  |  | .97 (.33)<sup>b</sup> | -2.1 [-6.36, 2.21] | -.32 [-.97, .33] | .027 |
|  |  |  |  |  | .62 (.44)<sup>c</sup> | -1.7 [-6.01, 2.65] | -.26 [-.91, .39] | .018 |
| Spo<sub>2</sub> (Free air) (%) | 83.4 ± 4.9 | 90.21 ± 3.4 | 84.4 ± 3.9 | 86.2 ± 4 | 10.63 (< .01)<sup>a</sup> | 3.99 [1.5, 6.47] | 1.07 [.37, 1.76] | .233 |
|  |  |  |  |  | 14.2 (< .01)<sup>b</sup> | 4.43 [2.04, 6.83] | 1.25 [.53, 1.95] | .294 |
| Distance of 3MWT (m) | 46 ± 24.3 | 149.1 ± 63 | 50 ± 24.4 | 67.3 ± 33.1 | 24.03 (< .01)<sup>a</sup> | 81.82 [47.93, 115.7] | 1.61 [.86, 2.35] | .407 |
|  |  |  |  |  | 67.9 (< .01)<sup>b</sup> | 91.44 [68.88, 113.99] | 2.72 [1.81, 3.62] | .666 |
|  |  |  |  |  | 64.33 (< .01)<sup>c</sup> | 91.62 [68.38, 114.86] | 2.67 [1.76, 3.55] | .661 |
| Level of Dyspnea | 5 ± 2 | 1.54 ± 1.2 | 5.19 ± 1.4 | 3.6 ± 1.5 | 22.89 (< .01)<sup>a</sup> | -2.08 [-2.96, -1.19] | -1.57 [-2.31, -.82] | .395 |
|  |  |  |  |  | 38.44 (< .01)<sup>b</sup> | -1.98 [-2.63, -1.33] | -2.04 [-2.83, -1.23] | .531 |
|  |  |  |  |  | 36.54 (< .01)<sup>c</sup> | -1.99 [-2.66, -1.32] | -2 [-2.79, -1.2] | .525 |
| RPE after Walking | 16.2 ± 1.6 | 10.8 ± 3 | 17 ± 1.3 | 14.8 ± 2 | 22.6 (< .01)<sup>a</sup> | -4.04 [-5.76, -2.32] | -1.56 [-2.3, -.82] | .393 |
|  |  |  |  |  | 18 (< .01)<sup>b</sup> | -3.46 [-5.12, -1.8] | -1.42 [-2.14, -.69] | .346 |
|  |  |  |  |  | 15.67 (< .01)<sup>c</sup> | -3.32 [-5.03, -1.62] | -1.34 [-2.05, -.62] | .322 |
| PH from VBG | 7.42 ± .08 | 7.5 ± .07 | 7.44 ± .06 | 7.43 ± .1 | 5.11 (.03)<sup>a</sup> | .06 [.01, .12] | .74 [.71, 1.41] | .127 |
|  |  |  |  |  | 8.38 (< .01)<sup>b</sup> | .08 [.02, .13] | .96 [.27, 1.63] | .198 |
|  |  |  |  |  | 7.31 (< .01)<sup>c</sup> | .07 [.02, .13] | .9 [.22, 1.57] | .181 |

PPT: Pulmonary physiotherapy; pO<sub>2</sub>: Mixed venous O<sub>2</sub> pressure; pCO<sub>2</sub>: Mixed venous CO<sub>2</sub> pressure; Spo<sub>2</sub>: Oxygen saturation; 3MWT: Three-minute walk test; RPE: Rating of perceived exertion; VBG: Venous blood gas; MD: Mean difference; CI: Confidence interval; SMD: Standardized mean difference (based on Cohen's d test), ANOVA: Analysis of variance, ANCOVA: Analysis of covariance.
<sup>a</sup> Expressed as mean ± standard deviation. The baseline and after values present the result of participants' assessment before and after three days, respectively.
<sup>\*\*</sup> Analyzed using ANOVA/ANCOVA tests. F statistics and P value were reported based on Group source analysis.
<sup>a</sup> Crude analysis
<sup>b</sup> Adjusted to baseline measurement of the variable (as covariance)
<sup>c</sup> Adjusted to baseline measurement of the variable and baseline O<sub>2</sub> saturation (free air) (as covariance)

Table 3 presents all of the SF-36 domains and summary scores after one month and three months. All scores were higher in the PPT group than the basic care group, except body pain after one month and role limitation due to physical health, mental health, and mental component summary score after three months. Only the differences in body pain domain score reach a statistically significant level.

**Table3.**
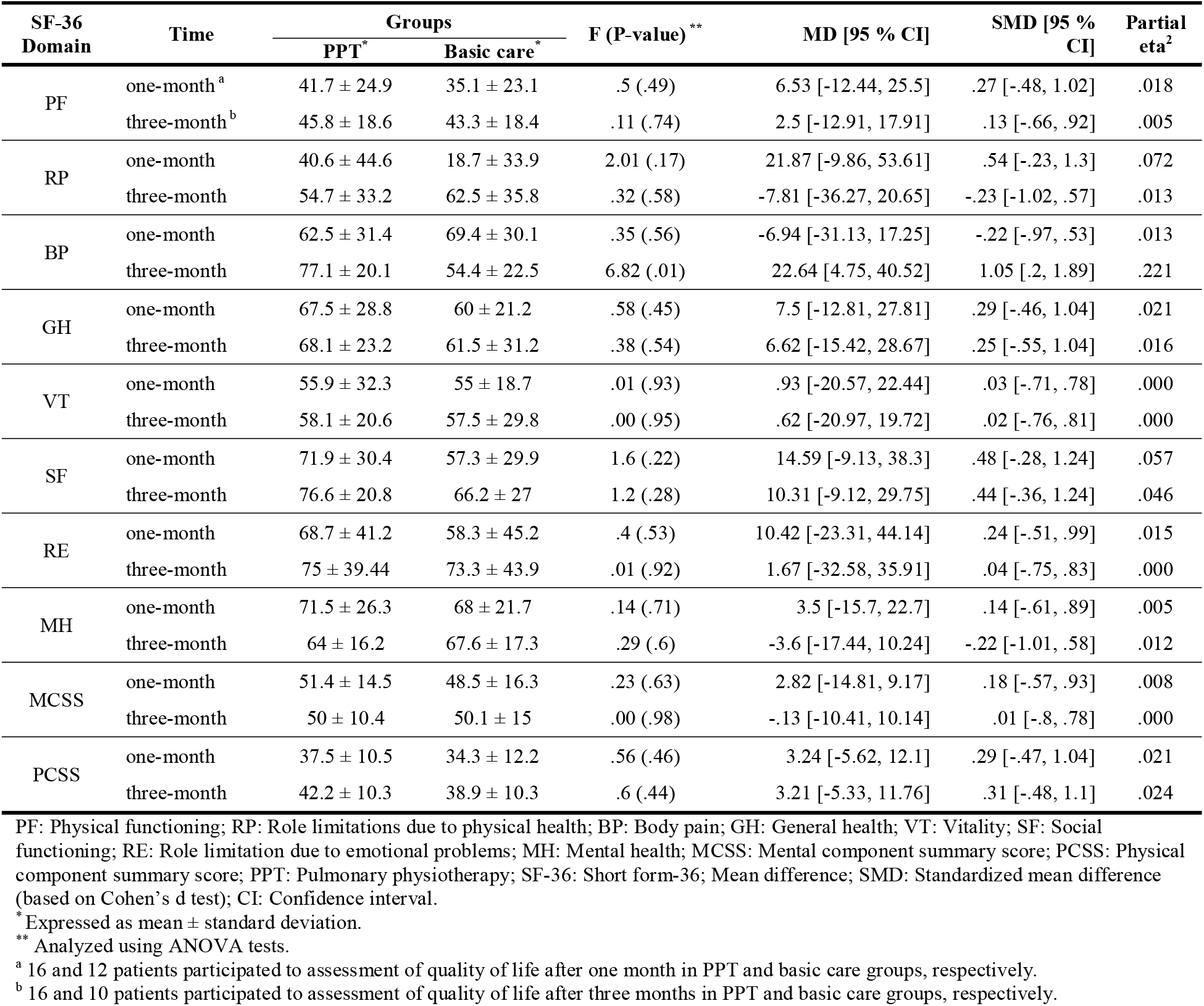
Distribution of SF-36 domains according to two arms in addition to related effect sizes after one and three months.

During one month after end of the intervention, two and six participants in the PPT group and basic care group were died (OR=.26 [.04, 1.49], P=.13, table 4). After considering baseline Spo2 as a covariate, OR was reduced (OR=.19 [.03, 1.30], P=.09). After three months, all allocated participants to PPT group were survived and one patient in basic care group was died (crude analysis: OR=.21 [.04, 1.16], P=.073; after considering baseline Spo2 as a covariate: OR=.16 [.26, 1.05], P=.056; table 4).

**Table 4.**
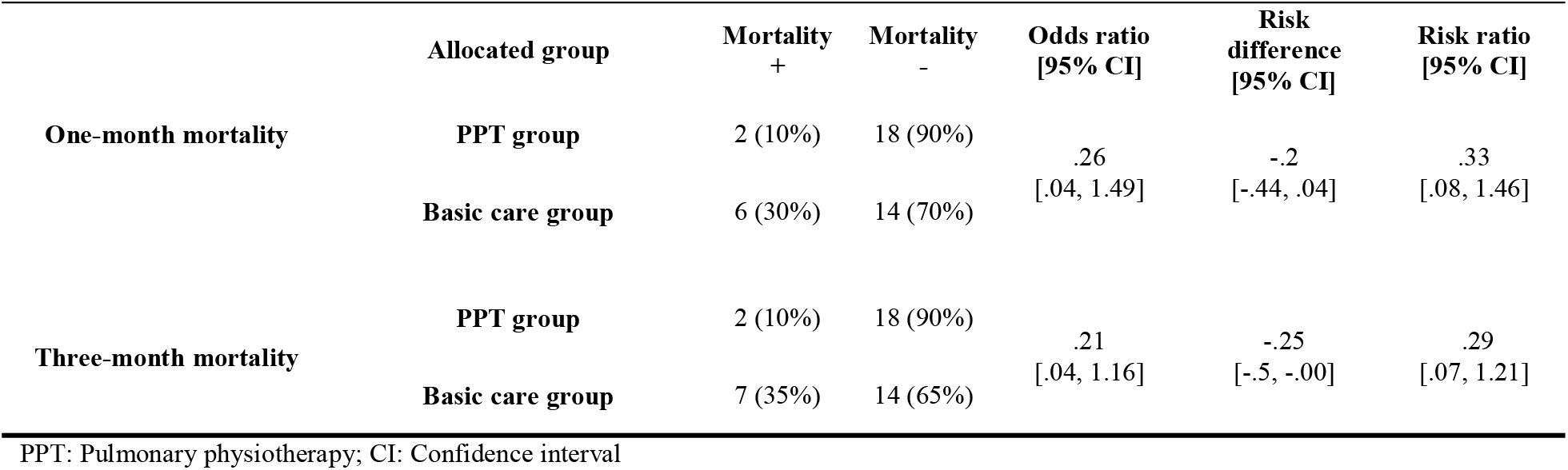
The mortality rate of participants allocated to groups in addition to related effect sizes.

No related adverse events were observed during the intervention period. The physiotherapists reported a minimal Spo2 decrease and minimal hemodynamic instability in some patients during PPT that was resolved a few minutes after the session. Also, the data monitoring committee confirmed that the reason for one case intubation in the experimental group during three days seems to not be related to the PPT.

## Discussion

According to our knowledge, this study is the first RCT to evaluate the efficacy of the PPT through comparison with a control group on non-intubated hospitalized patients with severe COVID-19 pneumonia. Our designed interventions seem to be safe and effective without serious adverse effects.

In general, the results show that participants’ pO2, PH, 3MWT, and level of dyspnea values were improved in the PPT group than the basic care group after adjustment of each measurement to the baseline measure. Spo2 was also higher in the PPT group than the basic care group clinically significant after three days [30]. Based on the result of standardized mean differences, PPT decreased pCO2 with a small effect size (NNT≈ 6) and improved pO2 (NNT≈ 2), venous blood PH (NNT= 2), level of dyspnea (NNT≈ 2), and RPE after walking (NNT≈ 2) with a large effect size, but inconclusive. PPT also improved Spo2 (NNT≈ 2) and distance of 3MWT (NNT≈ 2) with a very large effect size, but inconclusive. The NNT analyzes showed that it seems one patient with severe COVID-19 pneumonia can benefit from the PPT regarding Spo2, VBG analysis, functional capacity, and dyspnea measures if it is applied to at least two patients. After one-month follow-up, PPT was associated with higher HRQOL in role limitation due to physical health domain (NNT≈ 3) with a moderate effect size. Also, patients in PPT group presented higher score in body pain domain of SF-36 instrument than basic care group (NNT≈ 2) after three months with a large effect size. Patients with COVID-19 suffer from significant physical and psychological impairments that impact their HRQOL; Therefore, interventions improving HRQOL like rehabilitation should be considered in their management [36]. Although some limited domains of SF-36 presented higher value in the PPT group than the control group at one-month and three-month with very small effect size, but higher values of other domains in the PPT group than control group can suggest that prolonged duration rehabilitation, especially after discharge, may improve physical and psychological impairments and HRQOL. It has been shown that six weeks of respiratory rehabilitation could improve respiratory function, aerobic capacity, HRQOL, and anxiety of elderly patients with COVID-19, as reported by Liu and colleagues [14].

Allocated participants to the PPT group had a 20% and 25% lower one-month and three-month mortality rate than the basic care group, respectively. The NNT analysis shows that PPT kept alive one out of five patients until one month. However, the results of logistic test of one-month mortality rate is not statistically significant. Based on the NNT analysis, PPT can keep alive one out of four patients until three months. A similar finding was reported in patients after lobectomy after lung cancer by Çınar et al. (2020). They showed that patient received postoperative respiratory physiotherapy had no mortality after 30 days, while patients received standard postoperative care had 8% 30-day mortality rate [37]. In a systematic review and meta-analysis, Ryrsø et al. (2018) reported that supervised, early pulmonary rehabilitation could be associated to statistically significant mortality reduction (RR .58 [95% CI .34, .98]) in patients with acute exacerbation of chronic obstructive pulmonary disease compared to usual post-exacerbation care or no pulmonary rehabilitation program [38].

Adjusting each variable to its baseline measurement increased partial eta2> 10% than crude analysis, suggesting that even small differences of each measured outcome can change the results. A similar effect was not found about baseline Spo2.

There are currently some published studies with different levels of evidence, mostly review of investigations about PPT and other cases of viral pneumonia such as influenza, expert opinions, and consensus statements recommended how and when to use PPT for patients with different stages of COVID-19 pneumonia. Some researchers, physiotherapy, and rehabilitation experts have recommended PPT as an effective rehabilitative intervention for patients with all stages of COVID-19 pneumonia [4, 7, 11, 12]. Other researchers believed that only ACT should be applied in the presence of EPS to patients with the severe stage under special conditions; However, they have not suggested utilizing other PPT techniques like diaphragmatic breathing, respiratory muscle training, incentive spirometers, manual mobilization, and exercise training [1, 8, 9, 13]. They explained that acute patients often have a shallow and rapid respiratory pattern that has been adapted spontaneously to make the minimal effort; therefore, any change in this strategy may harm the patient. No other reason was given for considering PPT contraindicated in the severe stage of COVID-19 pneumonia [10]. We observed some of these events like Spo2 decreasing and cough stimulating, but all were resolved just some minutes after the session. In our clinical setting, we also sometimes observed that Spo2 decreases during breathing through continuous positive airway pressure, while using this non-invasive ventilator is recommended [39]. Our results showed that PPT seems not to be a contraindication for severe stage COVID-19, but it also may help therapeutic procedures.

About 34% of patients with COVID-19 pneumonia suffer from EPS [3]; therefore, ACT can be an effective treatment for them [14]. On the other hand, the main cell target of SARS-Cov-2 is type II pneumocyte, which is responsible for pulmonary surfactant synthesis [40]. As the pulmonary surfactant increases pulmonary compliance and facilitates the recruitment of collapsed airways [41], SARS-Cov-2 may cause alveolar stiffness. As there are some time-dependent properties of the tissue like creep and stress relaxation, a possible reason for the rationality of breathing exercises in the early phase can be their roles in tissue stiffness reduction or prevent pneumonia progression. Based on this hypothesis, early PPT in the acute phase may have therapeutic, preventive, and rehabilitative roles in the management of COVID-19 pneumonia. We did not have any assessment of lung tissue stiffness; therefore, our results support this possible hypothesis indirectly. Future researches with radiologic assessments that show tissue characteristics like chest computed tomography scan can test this hypothesis to some extent.

Our used PPT program had three main characteristics. First, choose the type of PPT technique(s) based on the patients’ signs and symptoms. We used IHT with emphasis on diaphragmatic breathing and educated them on how to cough and breathe properly, but ACTs were only applied to patients with EPS signs and symptoms. Second, we also defined some criteria for holding the intervention to get better safety. Third, we set a mild aerobic exercise in addition to chest physiotherapy to prevent muscle wasting and aerobic deterioration, which are seen in patients with COVID-19 [4]. We highly recommend that patients take a sufficient dose of oxygen and be under accurate monitoring during exercises.

Our research had some limitations which should be considered in future studies. First, we selected the sample size as MCID of Spo2 as the primary outcome measurement, but this number seems to have not enough power for other variables. For example, allocated participants in the PPT group had a 20% lower mortality rate than the control group. This RD seems to be influential; however, the results of logistic tests in the mortality rate did not reach a statistically significant level. Therefore, further investigations with a larger sample size are highly suggested evaluating safety and efficacy of PPT in patients with COVID-19. Second, we only recruited participants from a single center. Now, different centers have particular therapeutic guidelines that can impact the results of PPT. The applied interventions in this study with three-day duration, no need for specific tools and, low risk of adverse events suggest the feasibility of multi-center investigations on PPT effectiveness and safety on hospitalized patients with COVID-19. Third, we only recruited patients with a severe stage of COVID-19. Based on the mentioned possible mechanism, PPT may be more effective if to be started before disease severity reaches to severe phase even when the symptoms appear; therefore, future researchers are recommended to investigate the efficacy of PPT in patients with mild and moderate stages of COVID-19. Fourth, as participants of this study were from different cities, we could not re-assess their respiratory functions, aerobic capacity, and level of dyspnea. It is highly recommended that future researchers follow the objective pulmonary functions up to after patients’ discharge after applying PPT during the hospital stay.

## Conclusion

The results of this RCT suggest early PPT during three days, including airway clearance techniques, in the presence of excessive pulmonary secretion, and inspiratory hold breathing with emphasis to diaphragmatic breathing for hospitalized adult patients with a severe stage of COVID-19 pneumonia. Despite some recommendation, PPT seems to be safe and effective in the management of patients with severe COVID-19. The impact of early PPT on the mortality rate of these patients is currently indeterminate. Further researches with larger sample size are needed to evaluate the efficacy of PPT on COVID-19 patients with different severity stages.

## Data Availability

All individual participant data that underline the results reported in this article, after de-identification (text, tables, figures, and appendices), study protocol, statistical analysis plan and Informed consent form are available from the corresponding author and Mohammad Javaherian for researchers who provide a related approved proposal and ethical committee approve to achieve aims. Patient-level information will be anonymised and study documents will be redacted to protect the privacy of trial patients. These data will be available with publication date until one year.

## Contribution

MJ, AS, AK, MTB, ND, and BAM made substantial contributions to the conception and designed the study. MJ, ND, and SA contributed to applying the intervention or acquisition of the data. MJ and AK analysed the data. AK was responsible for the statistical analysis. BAM was responsible for data safety monitoring during the study. MJ, ND, and BAM interpreted the data and drafted the manuscript. MJ and BAM verified the data. All authors had access to the data, revised the manuscript, and approved it.

## Data sharing

All individual participant data that underline the results reported in this article, after de-identification (text, tables, figures, and appendices), statistical analysis plan and Informed consent form are available from the corresponding author and Mohammad Javaherian for researchers who provide a related approved proposal and ethical committee approve to achieve aims. Patient-level information will be anonymised and study documents will be redacted to protect the privacy of trial patients. These data will be available with publication date until one year.

## Acknowledgment

The researchers sincerely thank all physicians and nurses of intermediate care wards of IKHC. The authors would like to appreciate the methodologist’s support and constructive comments (s) research development office, IKHC, Tehran, Iran.

## Supporting information

**S1 Table.** The reason for protocol modifications.

| Type of Modification | Time of modification during the study | Reason(s) |
| --- | --- | --- |
| Changing the "level of dyspnea" from primary to secondary outcome measurement | Before enrollment | Their role in the sample size calculation. The role of these outcomes were not important in this research to be considered as primary outcome measurement. |
| Changing the "rating perceived of exertion after the three-minute walk test" from primary to secondary outcome measurement |  |  |
| Changing the "Venous Blood PH" from primary to secondary outcome measurement |  |  |
| Consider the history of intubation due to COVID-19 pneumonia as exclusion criteria. | Before enrollment | When the research team was preparing to start enrolling the participants, they found that there are cases weaned from mechanical ventilator recently. Based on defined inclusion/exclusion criteria, they could include in the study because they could consider patients with severe COVID-19. As the main aim of this study was to evaluate the efficacy and safety of pulmonary physiotherapy in the severe stage of COVID-19 to determine that can this treatment improve patients with COVID-19 and prevent their deterioration? Finally, the research team decided to consider the history of intubation due to COVID-19 as exclusion criteria. |

The permissions of all of the modifications were taken from the ethical committee of Tehran University of Medical Sciences.

